# Can ChatGPT-4o really pass medical science exams? A pragmatic analysis using novel questions

**DOI:** 10.1101/2024.06.29.24309595

**Authors:** Philip M. Newton, Christopher J. Summers, Uzman Zaheer, Maira Xiromeriti, Jemima R. Stokes, Jaskaran Singh Bhangu, Elis G. Roome, Alanna Roberts-Phillips, Darius Mazaheri-Asadi, Cameron D. Jones, Stuart Hughes, Dominic Gilbert, Ewan Jones, Keioni Essex, Emily C. Ellis, Ross Davey, Adrienne A. Cox, Jessica A. Bassett

## Abstract

ChatGPT apparently shows excellent performance on high level professional exams such as those involved in medical assessment and licensing. This has raised concerns that ChatGPT could be used for academic misconduct, especially in unproctored online exams. However, ChatGPT has also shown weaker performance on questions with pictures, and there have been concerns that ChatGPT’s performance may be artificially inflated by the public nature of the sample questions tested, meaning they likely formed part of the training materials for ChatGPT. This led to suggestions that cheating could be mitigated by using novel questions for every sitting of an exam and making extensive use of picture-based questions. These approaches remain untested.

Here we tested the performance of ChatGPT-4o on existing medical licensing exams in the UK and USA, and on novel questions based on those exams.

ChatGPT-4o scored 94% on the United Kingdom Medical Licensing Exam Applied Knowledge Test, and 89.9% on the United States Medical Licensing Exam Step 1. Performance was not diminished when the questions were rewritten into novel versions, or on completely novel questions which were not based on any existing questions. ChatGPT did show a slightly reduced performance on questions containing images, particularly when the answer options were added to an image as text labels.

These data demonstrate that the performance of ChatGPT continues to improve and that online unproctored exams are an invalid form of assessment of the foundational knowledge needed for higher order learning.

## Introduction

New generative artificial intelligence (AI) tools such as ChatGPT have attracted enormous attention, in part for their apparent ability to pass high level professional exams, with the subscription version of ChatGPT, running GPT-4, scoring an average of 75% on MCQ-based exams across a variety of disciplines (1). This excellent performance is replicated on specific medical qualifying exams such as the United States Medical Licensing Exam (USMLE) Step 1 where it scored 86% (2) and the United Kingdom Medical Licensing Exam Applied Knowledge Test (UK MLA AKT) where it scored 76.3% (3). These exams test high level problem-solving, requiring the application of core knowledge to clinical scenarios (4) and represent a broader principle wherein multiple choice questions can, if written appropriately, assess higher-order learning in a range of disciplines (5).

However there have been a number of responses and criticisms of the claim that ChatGPT is genuinely solving the problems presented in these questions, in part because this seems to lead logically onto the idea that ChatGPT is able to ‘reason’ which apparently it cannot (6). Instead, critics propose, tools like ChatGPT are more likely ‘regurgitating’ content which has been in their training materials (7), a proposal which is supported by the fact that many studies use sample papers which are in the public domain and have been for some time. For instance, the USMLE sample paper cited above was published in 2021. This regurgitation is not proposed to be verbatim, but instead is, essentially, a paraphrasing of prior training materials in a way that resembles a student who is plagiarising a piece of text by changing key words but without understanding the meaning, and so occasionally getting things (very) wrong (8). Thus, the argument goes, part of the reason why LLMs can ‘pass’ exams is because of this ‘regurgitation’ of sample papers which have been in the public domain for some time, and so to counter the apparent threat of ChatGPT to exam security and integrity educators could use novel questions for each sitting of the exam (9). In addition, there have been efforts to identify features of exam questions which ChatGPT might struggle with, for example an increase in the number of answer items, increasing language complexity or having multiple correct answers. However none of these appears to have any effect on the numbers of questions which ChatGPT can answer correctly (10).

Many early papers which tested the performance of ChatGPT on sample exams deliberately excluded questions containing images, on the basis that older versions of ChatGPT, even GPT-4, could not process these images. Thus, the reported performance of ChatGPT may be an over-estimation, since the percentage scored by ChatGPT uses a lower denominator once image-based questions are excluded (e.g. (11)). This also leads to proposals that educators could author ‘ChatGPT-proof’ questions by including images, along with mathematical calculations and reasoning tests, which it is proposed that ChatGPT does not perform well at (6).

These issues are important in part because of wider questions about the security, but also the inclusivity and cost, of examinations. In particular the sorts of university-administered knowledge tests that form part of a STEM curriculum prior to assessment using formal licensing examinations. Online examinations are cheaper and more flexible than their in-person equivalents, but they potentially increase the risk of cheating. During the COVID-19 pandemic, the percentage of students who admitted to cheating in online exams appeared to double, and more students reported cheating than not (12). One apparent solution to this problem is to increase the use of online proctoring/invigilation systems to monitor student behaviour. However, these then drive back up the cost of the online exams, and the student experience of remote proctoring is poor, with concerns about privacy, fairness, inclusivity and cost (13,14). An alternative is to avoid the use of proctoring altogether. A high profile 2023 publication analysed exam performance data from the COVID lockdown and concluded that unproctored online exams are a ‘valid and meaningful’ way of measuring student learning (15), although this analysis has been challenged (16) and does not include a consideration of ChatGPT. Thus it is important to understand whether ChatGPT truly can pass exams, including novel questions with images, as part of a consideration about how best to deploy exams, online or in-person, proctored or not.

Pragmatism is a research paradigm which prioritises the asking of questions whose answers will be useful, rather than perhaps asking more academic or basic questions (17). If ChatGPT truly can pass high level STEM exams, even with novel questions containing images, then from a pragmatic standpoint this is important because it essentially settles any debate about whether these examinations can be conducted in an online, unproctored format. From the pragmatic perspective, it does not matter *how* ChatGPT is doing this, either by truly solving problems or through some sophisticated paraphrasing. There is a related pragmatic issue, which is that for most STEM subjects there is a core curriculum; a basic set of knowledge and skills which graduates must be able to demonstrate in order to graduate, and also to be able to apply knowledge to practice. This cumulative view of learning has a long history and remains prevalent today through the use of instruments such as Bloom’s Taxonomy (18). In essence, we cannot expect students to undertake learning and practice at the higher levels of Blooms Taxonomy unless they have the core foundational knowledge to be applied to those higher levels. Thus educators need to assess that foundational knowledge first, before it is applied, particularly where there are safety concerns, e.g. for patients. However, it seems reasonable to propose that there are only so many ways that one can phrase any exam questions which might assess these core principles. This then creates a risk that, if educators strive to write completely novel questions on every core topic for every exam sitting, just to thwart ChatGPT, then this will rapidly become impossible. These issues also have relevance for the proposed positive benefits of ChatGPT. It offers great promise as a tutoring tool for students who are preparing for exams (19) but educators and learners both need to be confident that the answers given are logical and reasonable (20).

Some of the controversy and discourse about the apparent ability of ChatGPT to pass and perform well (or not) on exams likely comes from the frequent updating of ChatGPT over a short timescale. A review of ChatGPT’s performance on exams from multiple disciplines found that the subscription version of ChatGPT, running GPT-4, outperformed the free version running GPT-3 or 3.5, with the average difference being 25 percentage points (1). On May 13 2024 OpenAI, the creators of ChatGPT, released another update, entitled ChatGPT-4o, showing enhanced performance compared to GPT-4, particularly on the integration of text, visual and audio information (21). The performance of ChatGPT-4o on medical licensing exams has not yet been examined.

Here then we address the following research questions. It is important to be clear that the specific medical licensing-type exams tested here are intended to be a model for STEM exams generally, given that they are written to a high standard and are aimed at problem-solving and the application of knowledge (4,5).

1. How well does ChatGPT-4o perform on sample medical licensing exams in the USA and UK?
2. Is the performance of ChatGPT affected when these sample questions are rewritten into novel formats, but assessing the same core curricular concepts?
3. How well does ChatGPT perform on completely novel medical-licensing type questions?

## Methods

The following question sources were tested.

1. (Pilot) Wikiversity Fundamentals of Neuroscience Exam (22)
2. Sample paper 1, UK Medical Licensing Assessment Applied Knowledge Test (23)
3. USMLE Step 1 Sample paper (24)
4. Rewritten questions from 2+3
5. Completely Novel USMLE-style questions.

### Rewriting of existing questions in the public domain

Each question from sources 1-3 was rewritten by a member of the research team. Each question was rewritten three times with each rewrite undertaken by a different team member. Rewriting instructions were to create an original question, but which assessed the same learning, specifically to ‘change as much as possible about the question without changing the underlying learning. Change all the text where possible’. Suggestions of specific items to change included demographic details in the scenarios, answer options and answer order. Each team member was also provided with a summary of common issues found when writing USMLE-style questions (4) and asked to avoid any of the identified writing flaws. All rewritten items were checked for accuracy and originality by registered doctors (CS, RD) or a subject matter expert (PMN) and adjusted where necessary, for example if the revised question could be made even more different to the original question.

An initial pilot was undertaken using five questions on neuroscience from the ‘Wikiversity’ website. These were considered ‘lower order’ questions, assessing basic factual knowledge of neurological disease. The questions have been in the public domain since 2013. Each question was rewritten into three different forms by a member of the research team, who then discussed the process and feasibility of scaling the methodology to a larger exam. All four versions of each question were then pilot tested using GPT-4 on 23/04/24 and 24/04/24.

### Analysis of existing medical licensing exams and rewrites

Each question was tested using a single shot method in a way that would be expected to be the most likely approach taken by a student who was seeking to cheat on an MCQ exam, i.e. the text was highlighted in the pdf (original questions) or word document (rewrites), copied and then pasted directly into ChatGPT-4o with no attempt to format the text. Where the question included a picture, this was copied using screen clipping, saved and uploaded as a .png file with only the country and the question number as the file name (e.g. ‘UK32’). No additional prompts were given apart from the content of the question. Each question was asked in a new chat and no memory functions were activated. For the USMLE questions, a ‘temporary chat’ was activated for each question. No responses were given to ChatGPT. ChatGPT’s first response was recorded each time as correct/incorrect. ChatGPT-4o tests were undertaken May 14-24 2024.

### Creation and analysis of novel questions

Two sets of completely novel questions were generated, totalling 90 questions in all. A first set of forty novel questions were created in the style of questions for the UK MLA AKT and USMLE, by an author who is experienced in the creation of these assessment items (CS), according to guidance from the United States National Board of Medical Examiners (4). Ten of these questions included novel images that were either created for this study or were images from the private collection of one of the authors (CS). None of these images are available in the public domain. All images were obtained with appropriate consent and anonymised prior to use in keeping with paragraph 10 of the General Medical Council’s professional standards on making and using visual recordings of patients (25). These questions were mapped to curricula items from the MLA content map (26) and were of a comparative style and difficulty to the MLA. A second set of questions was written by an author (PMN) using guidance for the creation of multiple-choice questions which assess higher order learning in STEM. These guidelines include identifying assumed knowledge, creating problem-solving scenarios and the use of actions as answer options (5). Some of these questions included images sourced from Wikimedia Commons. During this process the authors observed a trend that ChatGPT appeared to struggle with anatomical images that had novel text labels, e.g. a brain section with the labels A-H added, with arrows to specific brain regions that corresponded to question answers. To probe this further, an additional set of questions was generated so that there was a total of 14 pairs of questions which assessed the same learning but either using a labelled image, or text equivalent. Finally, ChatGPT was then asked simply to identify the labels on the images from these questions where possible. Each question was asked in a new ‘temporary chat’. ChatGPT-4o tests were undertaken May 24-Jun 18 2024.

## Results

### Summary

We tested a total of 705 assessment items, of which ChatGPT answered 635 (90%) correctly. 111 of these questions contained images, of which ChatGPT answered 76 (68.5%) correctly. A breakdown of these items is below.

### Wikiversity Pilot

GPT-4 correctly answered all versions of all questions, both the originals and the rewritten versions.

### United Kingdom Medical Licensing Assessment, Applied Knowledge Test

ChatGPT-4o answered 94 of 100 questions on the original sample paper. Five of the questions included pictures. ChatGPT answered four of these correctly. ChatGPT then scored 93%, 91% and 95% on the three collections of rewrites. One question, on herpes zoster ophthalmicus, was answered incorrectly on all four occasions. In all other cases there was no consistent pattern. Some questions that ChatGPT had answered incorrectly on the original sample paper were answered correctly once rewritten, but the converse was also true for other questions. 85% of questions were answered correctly in all four versions (original and all three rewrites). The full dataset and questions are in Supplementary Data S1.

### United States Medical Licensing Exam Step 1

ChatGPT-4o scored 89.9% (107/119) of the original questions correctly. Of the original 119, there were images in 23 of them, of which 16 (69.6%) were answered correctly. This suggested that ChatGPT might struggle more with the picture questions in this exam. Given that ChatGPT-4o had already demonstrated no impairment of performance when rewriting text questions from the UK MLA AKT into a novel format, we decided to rewrite only a sample of 27 of the USMLE questions, but to probe further this possible diminished performance on questions containing pictures by including 13 picture questions, of which 5 had been answered incorrectly from the original paper. Of the sample of 27, ChatGPT scored 74.1% (20/27) on the original versions, and then 85.2 (23/27), 70.4% (19/27) and 85.2% (23/27) on the rewrites. Only one question was answered incorrectly in all four versions. This was a picture question based on a graph, while the other four picture questions which ChatGPT had answered incorrectly were then answered correctly at least once during the rewrites. 55.6% (15/27) of questions were answered correctly on all four occasions. The full dataset and questions are in Supplementary Data S1.

### Novel questions

A total of 90 novel questions were generated, of which ChatGPT answered 75 correctly. 28 of these questions were in pairs (2×14) which assessed the same learning in each pair. One version of the question contained a labelled image where the labels were simple letters (A,B,C etc) and these were the answer options, for example the image was a picture of the brain with different regions labelled A-H. The paired question contained answer options in text form, for example the brain regions were listed as text. An example of this format is in Figure 1. ChatGPT answered 13/14 of the text version of these questions, but only 2/14 of the labelled image questions. A summary of the analysis is in Supplementary Data S1. The novel questions may be shared upon request but are not published here due to the images contained within.

**Figure 1.**
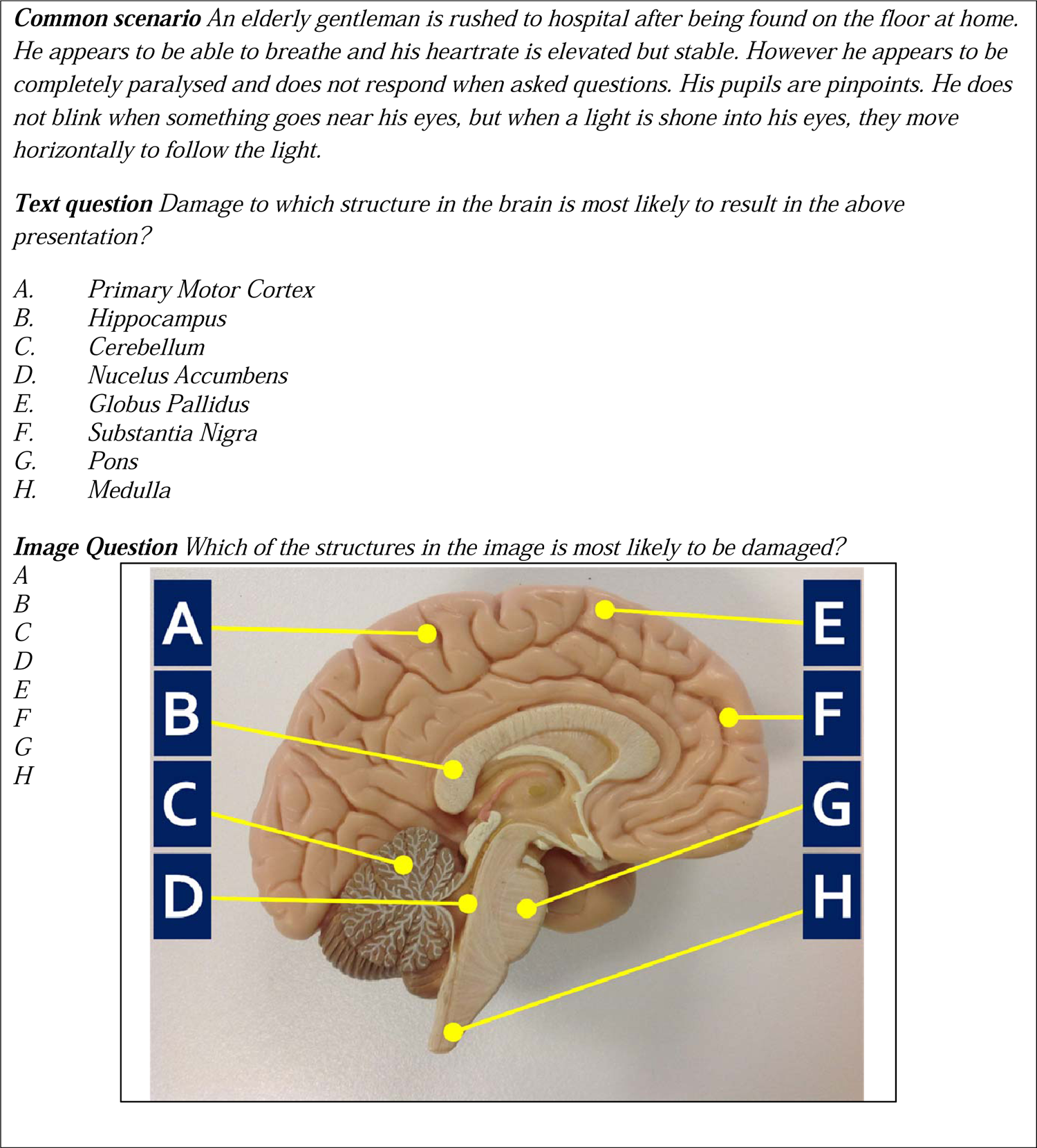
An example of a novel-higher order MCQ written using established guidelines (5), with text options as answers (which ChatGPT answers correctly), or a labelled image (which ChatGPT answers incorrectly). Note that the answer options do not correspond exactly.

### Identification of labels on images

Ten of the labelled images were structured in a way that it was reasonable to upload them to ChatGPT-4o with the prompt ‘Can you identify all the labels (A-X) on the uploaded image?’ where ‘X’ was either E, F, G or H depending on the number of labels. Of a total of 66 labels across the 10 images, ChatGPT correctly labelled 25 items. For all 10 images ChatGPT correctly identified the main structure in the image (e.g. brain, kidney) but not the labelled subregions.

## Discussion

ChatGPT-4o showed a very high level of performance on the papers tested, even when the questions were rewritten so that they assessed the same learning but with different wording. This level of performance was also found on completely novel questions written in the style of professional licensing exams. Our analysis included many questions based on images, and almost all questions were designed to assess higher-order problem-solving (4,5).

A repeated finding from the research on academic misconduct demonstrates that one of the strongest factors contributing to an increased likelihood in the occurrence of academic dishonesty is the ease with which it can be committed (12,27). Cheating in online exams was already high before the emergence of ChatGPT (12) and our findings demonstrate that any student using ChatGPT would likely receive an excellent mark even if they had no prior knowledge whatsoever, further increasing any temptation to cheat. Thus it seems reasonable to propose that our findings mean online unproctored summative exams are now no longer a valid form of assessment, a conclusion which is in contrast to findings published following an analysis of exam performance during the COVID pandemic, but before the emergence of ChatGPT (15).

The high performance levels of ChatGPT may also increase the temptation to cheat using ChatGPT even in proctored exams, particularly if they are taken online; data suggest that proctoring considerably reduces cheating in online exams but does not eliminate it completely (12). We are not aware of any current data on the extent to which students are using ChatGPT to cheat in online exams, proctored or unproctored, although this is the subject of ongoing work. A study conducted in Vietnam in May 2023 showed that 23.7% of undergraduates cheated using ChatGPT, although the assessment formats were not specified (28). A study conducted at around the same time in US high schools found similar numbers in one school, though lower in two others (29). These figures seem likely to increase as ChatGPT becomes better known and more widely available, along with similar tools such as Claude.AI.

One intuitive response to these challenges is to design questions which ChatGPT finds harder to answer. This ‘arms race’ approach is partly the genesis of the current paper, based in part on earlier studies which observed that ChatGPT could not process image-based questions at all, and other studies suggesting that ChatGPT is a ‘copy and paste’ machine whose impact can be minimized by using novel questions for each sitting of an exam (9). We did find that ChatGPT struggled more on a very specific type of MCQ, where the answer items were single letter labels and arrows on images. There is more than one possible explanation for this apparent weakness. These questions are designed to require ‘assumed knowledge’ and so to be harder to answer than factual recall questions (5). For example, the picture item shown in figure 1 requires the test taker to know that the scenario represents the clinical condition Locked-In Syndrome, and then to know that this condition is associated with damage to the part of the brain called the pons, and then to be able to identify the anatomical location of the pons on a picture of a model. ChatGPT consistently struggled with these specific types of image questions and so one interpretation is that it is the ‘multi-step’ nature of these questions which trips up ChatGPT. However, ChatGPT was consistently correct on the text versions of these questions and would give detailed descriptions of the answer option. ChatGPT was also clearly able to identify, in text form, where the pons is located (for example). But when simply asked to identify the labels on these images ChatGPT struggled, indicating that it is the processing of these specific types of text-labelled images which ChatGPT struggles with, rather than the solving of multi-step problems.

One intuitive conclusion from these findings with images is that such questions could be used to thwart ChatGPT and so deter cheating in online exams. However, we caution against this interpretation. Writing an entire exam based on these types of questions seems implausible and unlikely to be valid. This limitation likely applies to other methods identified as a way of ‘defeating’ ChatGPT. For example, an older study, using an unidentified version of ChatGPT, showed that ChatGPT overselects answer options ‘all of the above’ or ‘none of the above’, meaning that when these answer options are present but are incorrect, ChatGPT shows a much lower performance compared to when these answer options are absent or when they are present but are the correct answer. However, designing questions which incorporate this flaw also seems likely to be a short-term measure that may well result in poorer quality questions and weaker curriculum coverage. These types of answer options are also advised against when writing high quality assessment items (5).

Any reduction in the use of online unproctored exams will clearly not eradicate academic misconduct. There are a wide range of dishonest behaviours undertaken by medical and other students (30), and the performance of ChatGPT on assessment formats such as essays is also very strong (31). Essays are, by design, asynchronous and unmonitored, meaning that it would be almost impossible to prevent a student from using ChatGPT to complete assignments in these formats. Detection tools have been developed and these appear to show good accuracy for raw text generated by tools such as ChatGPT (32) but they can be easily circumvented (33) and even a very small rate of false-positives is problematic since there is no independent source to match a student assignment to, unlike with ‘conventional’ plagiarism, meaning that problematic, adversarial situations can quickly arise when students are accused of cheating on essays using ChatGPT (34).

The performance of ChatGPT-4o demonstrated here shows a modest improvement when compared to that seen using GPT-4, which itself shows a much improved performance compared to GPT-3 and GPT-3.5 (1), although many prior papers excluded image-based questions from their analyses whereas they are included here. This trend of improving performance seems likely to continue; at the time of writing (July 2024), OpenAI are rolling out enhanced visual recognition features in GPT-4o to their subscribers, meaning that users will be able to simply point their camera at an exam question and it will scan and ‘read’ the text before generating an answer (21).

The high performance of ChatGPT-4o on the exams tested here and elsewhere leads naturally to a question of whether these tools might also be able to *write* such exams. A review on some of the older versions of these tools concluded that question generation is possible although with some limitations, and proposed further testing (35). It is now possible to upload considerable volumes of data to ChatGPT and to build custom GPTs which have specific instructions tailored to certain tasks, as designed by the creator. This approach has already shown promise for the creation of USMLE-style assessment items and may even be able to generate an entire exam and blueprint it to a curriculum, saving considerable time and cost for educators and universities (36). This possibility arose during the conduct of the study here wherein some questions that were initially answered incorrectly by ChatGPT revealed either strong distractors or potential ambiguities in the question stem or associated image, suggesting weaknesses in the question itself. No questions tested here were eliminated from analysis for being actually incorrect or of poor quality, but this analysis suggested that such issues might be easily identified by using ChatGPT as an adjunct to exam creation and standard setting.

Similar benefits could also be obtained for students. The research team here noted the accuracy and value of the explanations provided by ChatGPT when answering the questions, and these naturally suggest the potential of ChatGPT, and the aforementioned custom GPTs, as study tools for students. Such an approach has been successfully used in ophthalmology (37) and anatomy learning (38).

## Conclusion

ChatGPT-4o shows very high levels of performance on MCQ-based applied knowledge tests, including questions with images. These data echo but improve further upon findings from earlier versions of ChatGPT (39) and suggest that educators will find it extremely difficult to write questions which are ‘ChatGPT-proof’, even if they are completely novel and image-based. The logical conclusion is that unproctored online exams are no longer a valid form of assessment, even when assessing higher order learning. These assessments, and lower-level MCQs based exams testing core foundational knowledge, should only be conducted under secure conditions.

## Data Availability

All data produced in the present work are contained in the manuscript

## References

1. Newton P, Xiromeriti M. ChatGPT performance on multiple choice question examinations in higher education. A pragmatic scoping review. Assess Eval High Educ. 2024;0(0):1–18.

2. Garabet R, Mackey BP, Cross J, Weingarten M. ChatGPT-4 Performance on USMLE Step 1 Style Questions and Its Implications for Medical Education: A Comparative Study Across Systems and Disciplines. Med Sci Educ. 2024 Feb 1;34(1):145–52.

3. Lai UH, Wu KS, Hsu TY, Kan JKC. Evaluating the performance of ChatGPT-4 on the United Kingdom Medical Licensing Assessment. Front Med. 2023 Sep 19;10:1240915.

4. Billings M, DeRuchie K, Hussie K, Kulesher A, Merrell J, Morales A, et al. Constructing written test questions for the Health Sciences [Internet]. National Board of Medical Examiners; 2020 [cited 2022 Apr 7]. Available from: https://www.nbme.org/sites/default/files/2020-11/NBME_Item%20Writing%20Guide_2020.pdf

5. Newton PM. Guidelines for Creating Online MCQ-Based Exams to Evaluate Higher Order Learning and Reduce Academic Misconduct. In: Eaton SE, editor. Handbook of Academic Integrity [Internet]. Singapore: Springer Nature; 2023 [cited 2023 Jul 13]. p. 1–17. Available from: 10.1007/978-981-287-079-7_93-1

6. Arkoudas K. GPT-4 Can’t Reason [Internet]. arXiv; 2023 [cited 2024 Feb 18]. Available from: http://arxiv.org/abs/2308.03762

7. Yeo YH, Samaan JS, Ng WH, Ting PS, Trivedi H, Vipani A, et al. Assessing the performance of ChatGPT in answering questions regarding cirrhosis and hepatocellular carcinoma. Clin Mol Hepatol. 2023 Jul;29(3):721–32.

8. Marcus G. Partial Regurgitation and how LLMs really… [Internet]. Marcus on AI. 2024 [cited 2024 Jun 3]. Available from: https://garymarcus.substack.com/p/partial-regurgitation-and-how-llms/comments

9. Lo CK. What Is the Impact of ChatGPT on Education? A Rapid Review of the Literature. Educ Sci. 2023 Apr;13(4):410.

10. Ram S, Qian C. A Study on the Vulnerability of Test Questions against ChatGPT-based Cheating. In: 2023 International Conference on Machine Learning and Applications (ICMLA) [Internet]. 2023 [cited 2024 Jun 17]. p. 1710–5. Available from: https://ieeexplore.ieee.org/abstract/document/10460039

11. Abbas A, Rehman MS, Rehman SS. Comparing the Performance of Popular Large Language Models on the National Board of Medical Examiners Sample Questions. Cureus. 16(3):e55991.

12. Newton PM, Essex K. How Common is Cheating in Online Exams and did it Increase During the COVID-19 Pandemic? A Systematic Review. J Acad Ethics [Internet]. 2023 Aug 4 [cited 2023 Aug 7]; Available from: 10.1007/s10805-023-09485-5

13. Marano E, Newton PM, Birch Z, Croombs M, Gilbert C, Draper MJ. What is the student experience of remote proctoring? A pragmatic scoping review. High Educ Q. n/a(n/a):e12506.

14. Meulmeester FL, Dubois EA, Krommenhoek-van Es C (Tineke), de Jong PGM, Langers AMJ. Medical Students’ Perspectives on Online Proctoring During Remote Digital Progress Test. Med Sci Educ. 2021 Sep 30;31(6):1773–7.

15. Chan JCK, Ahn D. Unproctored online exams provide meaningful assessment of student learning. Proc Natl Acad Sci. 2023 Aug;120(31):e2302020120.

16. Newton PM. The validity of unproctored online exams is undermined by cheating. Proc Natl Acad Sci. 2023 Oct 10;120(41):e2312978120.

17. Newton PM, Da Silva A, Berry S. The Case for Pragmatic Evidence-Based Higher Education: A Useful Way Forward? Front Educ [Internet]. 2020 [cited 2021 May 8];5. Available from: https://www.frontiersin.org/articles/10.3389/feduc.2020.583157/full

18. Newton PM, Da Silva A, Peters LG. A Pragmatic Master List of Action Verbs for Bloom’s Taxonomy. Front Educ [Internet]. 2020 [cited 2020 Jul 14];5. Available from: https://www.frontiersin.org/articles/10.3389/feduc.2020.00107/full

19. Koga S. The Potential of ChatGPT in Medical Education: Focusing on USMLE Preparation. Ann Biomed Eng. 2023 Oct 1;51(10):2123–4.

20. Daungsupawong H, Wiwanitkit V. ChatGPT-4 Performance on USMLE Step 1 Style Questions and Its Implications for Medical Education: Correspondence. Med Sci Educ [Internet]. 2024 Apr 5 [cited 2024 Jun 3]; Available from: 10.1007/s40670-024-02033-9

21. OpenAI. Hello GPT-4o [Internet]. [cited 2024 Jun 3]. Available from: https://openai.com/index/hello-gpt-4o/

22. Wikiversity. Fundamentals of Neuroscience/Exams - Wikiversity [Internet]. 2013 [cited 2024 Feb 10]. Available from: https://en.wikiversity.org/wiki/Fundamentals_of_Neuroscience/Exams

23. Medical Schools Council. Practice exam for the MS AKT | Medical Schools Council [Internet]. 2023 [cited 2024 Mar 10]. Available from: https://www.medschools.ac.uk/medical-licensing-assessment/preparing-for-the-ms-akt/practice-exam-for-the-ms-akt

24. United States Medical Licensing Examination. Step 1 Sample Test Questions | USMLE [Internet]. 2021 [cited 2024 Jun 10]. Available from: https://www.usmle.org/prepare-your-exam/step-1-materials/step-1-sample-test-questions

25. GMC. Making and using visual and audio recordings of patients (summary) [Internet]. General Medical Council; 2011 [cited 2023 Jun 15]. Available from: https://www.gmc-uk.org/professional-standards/professional-standards-for-doctors/making-and-using-visual-and-audio-recordings-of-patients

26. GMC. MLA content map [Internet]. 2021 [cited 2024 Jun 15]. Available from: https://www.gmc-uk.org/education/medical-licensing-assessment/mla-content-map

27. Bretag T, Harper R, Burton M, Ellis C, Newton P, Rozenberg P, et al. Contract cheating: a survey of Australian university students. Stud High Educ. 2019 Nov 2;44(11):1837–56.

28. Nguyen HM, Goto D. Unmasking academic cheating behavior in the artificial intelligence era: Evidence from Vietnamese undergraduates. Educ Inf Technol [Internet]. 2024 Feb 5 [cited 2024 Feb 18]; Available from: 10.1007/s10639-024-12495-4

29. Lee VR, Pope D, Miles S, Zárate RC. Cheating in the age of generative AI: A high school survey study of cheating behaviors before and after the release of ChatGPT. Comput Educ Artif Intell. 2024 Dec 1;7:100253.

30. Henning MA, Chen Y, Ram S, Malpas P. Describing the Attributional Nature of Academic Dishonesty. Med Sci Educ. 2019 Jun 1;29(2):577–81.

31. Herbold S, Hautli-Janisz A, Heuer U, Kikteva Z, Trautsch A. AI, write an essay for me: A large-scale comparison of human-written versus ChatGPT-generated essays [Internet]. arXiv; 2023 [cited 2023 May 8]. Available from: http://arxiv.org/abs/2304.14276

32. Weber-Wulff D, Anohina-Naumeca A, Bjelobaba S, Foltýnek T, Guerrero-Dib J, Popoola O, et al. Testing of Detection Tools for AI-Generated Text [Internet]. arXiv; 2023 [cited 2023 Aug 7]. Available from: http://arxiv.org/abs/2306.15666

33. Perkins M, Roe J, Vu BH, Postma D, Hickerson D, McGaughran J, et al. arXiv.org. 2024 [cited 2024 Jun 11]. GenAI Detection Tools, Adversarial Techniques and Implications for Inclusivity in Higher Education. Available from: https://arxiv.org/abs/2403.19148v1

34. Gorichanaz T. Accused: How students respond to allegations of using ChatGPT on assessments. Learn Res Pract [Internet]. 2023 Jul 3 [cited 2024 May 3]; Available from: https://www.tandfonline.com/doi/abs/10.1080/23735082.2023.2254787

35. Artsi Y, Sorin V, Konen E, Glicksberg BS, Nadkarni G, Klang E. Large language models for generating medical examinations: systematic review. BMC Med Educ. 2024 Mar 29;24(1):354.

36. Kıyak YS, Kononowicz AA. Case-based MCQ generator: A custom ChatGPT based on published prompts in the literature for automatic item generation. Med Teach [Internet]. 2024 Feb 6 [cited 2024 Jun 11]; Available from: https://www.tandfonline.com/doi/abs/10.1080/0142159X.2024.2314723

37. Sevgi M, Antaki F, Keane PA. Medical education with large language models in ophthalmology: custom instructions and enhanced retrieval capabilities. Br J Ophthalmol [Internet]. 2024 May 7 [cited 2024 Jun 11]; Available from: https://bjo.bmj.com/content/early/2024/05/07/bjo-2023-325046

38. Collins BR, Black EW, Rarey KE. Introducing AnatomyGPT: A customized artificial intelligence application for anatomical sciences education. Clin Anat [Internet]. [cited 2024 Jun 11];n/a(n/a). Available from: https://onlinelibrary.wiley.com/doi/abs/10.1002/ca.24178

39. Sood A, Mansoor N, Memmi C, Lynch M, Lynch J. Generative pretrained transformer-4, an artificial intelligence text predictive model, has a high capability for passing novel written radiology exam questions. Int J Comput Assist Radiol Surg. 2024 Apr 1;19(4):645–53.

